# Improved survival outcome in SARs-CoV-2 (COVID-19) Acute Respiratory Distress Syndrome patients with Tocilizumab administration

**DOI:** 10.1101/2020.05.13.20100081

**Authors:** Nafisa Wadud, Naim Ahmed, Mannu Shergil, Maida Khan, Murali Krishna, Aamir Gilani, Samer El Zarif, Jodi Galaydick, Karthika Linga, Shravan Koor, Julia Galea, Lauren Stuczynski, Maria B Osundele

## Abstract

**Background:** The novel human coronavirus, severe acute respiratory syndrome coronavirus-2 (SARs-CoV-2), was declared a global pandemic by the World Health Organization on March 11, 2020. Hence, there is an urgency to find effective treatment. Of those patients afflicted in the United States, many have required treatment with ventilator secondary to acute respiratory distress syndrome (ARDS). Data are needed regarding the benefit of treatment and prevention of the cytokine storms in COVID-19 patients with Tocilizumab.

**Methods:** Clinical outcomes data for patients admitted to Orange Regional Medical Center with confirmed COVID-19 from Mar 15, 2020 to Apr 20, 2020 were identified through electronic health record chart review. We conducted a retrospective case-control study in confirmed COVID 19 positive patients with ARDS requiring mechanical ventilation and compared outcome in terms of mortality and length of stay amongst those who received Tocilizumab as treatment modality opposed to those that did not.

**Results:** A total of 94 patients with COVID-19 ARDS were analyzed. 44 were in the study group and 50 in the control group. We tried to match both group as close as possible in terms of age, sex, BMI and HS score-calculated using inflammatory markers-ferritin, triglycerides, AST and fibrinogen. The median age was 55.5 years in the study group and 66 in the control group, difference was not statistically significant. Average HS score was 114 in the Tocilizumab group and 92 in the control group, difference was statistically significant with P<0.0001. Also, the patients in the study group had elevated levels of IL-6, triglycerides, AST, ferritin which were statistically significant with p < 0.0001 when compared to the control group. Length of stay was longer, average 17.9 days in the Tocilizumab. Survival rate was much lower at 48 % in the control group and 61.36 % in patients who received Tocilizumab with significant P value of < 0.00001. The number needed to treat (NNT) was 7.48, if we treat 8 patients with Tocilizumab, 1 will not die.

**Conclusions:** Cytokine Release Syndrome (CRS) occurs in a large number of patients with severe COVID-19, which is also an important cause of death. IL-6 is the key molecule of CRS, so IL-6R antagonist Tocilizumab may be of value in improving outcomes. In our study Tocilizumab group seemed to have improved survival outcome. Results have to be interpreted with caution since this is a retrospective study and mortality is affected by multiple, confounding factors. We await the results of ongoing randomized controlled trials to definitely answer the question of whether Tocilizumab improves survival in COVID-19 ARDS patients.

## INTRODUCTION

The novel human coronavirus, Severe acute respiratory syndrome coronavirus-2 (SARs-CoV-2), was declared a global pandemic by the World Health Organization on March 11, 2020. Over 2.7 million confirmed cases and 190,890 deaths have been identified worldwide^1^. In the USA, there have been 52,400 deaths as of April 24, 2020^1^, New York State being the epicenter of pandemic^2^. Hence, there is an urgency to find effective treatment. Of those patients afflicted in the United States, many have required treatment with ventilator secondary to acute respiratory distress syndrome.

The pathogenesis of severe acute respiratory syndrome (SARS) related to coronavirus involves a cytokine storm with high serum levels of pro-inflammatory cytokines and chemokines interleukin 6 (IL-6). The pro-inflammatory cytokine IL-6 seems to have a prominent role in this inflammatory cascade, which may result in increased alveolar-capillary blood-gas exchange dysfunction^3,4^. The most recent clinical experiences in China and Italy ^5^ suggested that patients admitted to intensive care units, the cytokine storm syndrome was proportional to the severity of disease^6^, often progressing to cardiovascular collapse, multiple organ dysfunction and death rapidly^7^. Therefore, early identification, treatment and prevention of the cytokine storms are of crucial importance for these patients.

Tocilizumab is a blocker of IL-6R, which can effectively block IL-6 signal transduction pathway. The safety of tocilizumab in phase III double-blind controlled trials was studied in patients with rheumatoid arthritis. There were no complications associated with tocilizumab and no history of illness deterioration or death. Overall, the risk of secondary infection with Tocilizumab is not too high^8^. We recommend treatment of hyperinflammation using existing, approved therapies with proven safety profiles to address the immediate need to reduce the rising mortality. Data are needed regarding the benefit of treatment and prevention of the cytokine storms in COVID-19 patients with Tocilizumab.

Currently, a small clinical trial in China has shown good efficacy in tocilizumab. After a few days of treatment, patients experienced clinical improvement, became afebrile and with gradually decreased oxygen consumption, all other symptoms were significantly improved. CT chest showed regression of the pulmonary infiltrates and ground glass appearance^9^. Laboratory examination showed improvement in peripheral blood lymphocytes and C-reactive protein, suggesting that tocilizumab could be an efficient treatment for COVID-19 patients. A multicenter, randomized controlled trial for the efficacy and safety of tocilizumab in the treatment of new coronavirus pneumonia (COVID-19 is underway.^10,11^

Secondary haemophagocytic lymphohistiocytosis (sHLH) is an under-recognized, characterized by a fulminant and fatal hypercytokinaemia with multiorgan failure^12^. A cytokine profile resembling sHLH is associated with COVID-19 disease severity. All patients with severe COVID-19 should be screened for hyperinflammation using laboratory trends (eg, increasing ferritin, decreasing platelet counts, or CRP) and the HScore to identify the subgroup of patients for whom immunosuppression could improve mortality. HScores greater than 169 are 93% sensitive and 86% specific for HLH^12^.

## METHODS

### Study Population

Orange Regional Medical Center serves racially and ethnically diverse patient population in the Hudson Valley. In this study, patient data came from ICU and floor during the Coronavirus pandemic. We included patients who were at least 18 years old, had a laboratory confirmed Covid-19 infection, and were admitted to the hospital between Mar 15, 2020 to Apr 20, 2020. A confirmed case of Covid-19 was defined by a positive reverse transcriptase polymerase chain reaction (RT-PCR) assay of a specimen collected via nasopharyngeal swab. The Orange Regional Medical Center Institutional Review Board approved this research under a regulatory protocol allowing for analysis of limited patient data.

### Data Collection

We obtained demographics, laboratory test results, diagnosis codes (International Classification of Diseases-10), procedures, ICU and floor progress notes during hospitalization. Demographics included age, sex, BMI, race, ethnic group in the electronic health records. All laboratory values (CRP, IL-6, TGL, ferritin, fibrinogen, AST, D-dimer/ fibrinogen, EKG) were obtained as part of indicated clinical care. Treatment modalities used were also compared (Tocilizumab - Dose in mg/kg, One or two doses, Hydroxychloroquine, Azithromycin, Steroids - hydrocortisone/ methylprednisolone/ dexamethasone). For patients who sustained acute kidney injury – mode of management – Hemodialysis, CRRT were also obtained. During ICU stay Ventilator setting: Average PEEP, Average FiO2, Average plateau pressure, Average driving pressure; use of Paralytic medication (cisatracurium, atracurium, rocuronium or vecuronium infusions) and proning were taken into account for data analysis.

### Definitions of Co-morbidities

We defined co-morbidities as the presence of diagnosis codes associated with asthma, COPD, Diabetes, use of immunosuppressant medications.

### Definitions of Outcomes

We assessed length of stay – number of days on ventilator, in-hospital and Intensive care unit; mortality, survival and discharge (home, rehab, transfer to outside facility) during admission.

### Statistical Analysis

We used descriptive statistics to summarize the data; results are reported as Medians, ranges, averages and p-values as appropriate. Categorical variables were summarized as counts and percentages. Compared by Z score and comparison of mean between Tocilizumab and non-Tocilizumab group within 1 Standard deviation and 95% Confidence Interval. All statistical tests were 2-tailed, and statistical significance was defined as *P* <0.05. https://www.medcalc.org/

## RESULTS

A total of 94 patients with COVID-19 ARDS were analyzed. 44 were in the study group and 50 in the control group. We tried to match both group as close as possible in terms of age, sex, BMI and HS score-calculated using inflammatory markers-ferritin, triglycerides, AST and fibrinogen. The median age was 55.5 years in the study group and 66 in the control group, difference was not statistically significant. Average HS score was 114 in the Tocilizumab group and 92 in the control group, difference was statistically significant with P<0.0001. Also, the patients in the study group had elevated levels of IL-6, triglycerides, AST, ferritin which were statistically significant with p < 0.0001when compared to the control group. Length of stay was longer, average 17.9 days in the Tocilizumab. Survival rate was much lower at 48 % in the control group and 61.36 % in patients who received Tocilizumab with significant P value of < 0.00001. The number needed to treat (NNT) was 7.48, if we treat 8 patients with Tocilizumab, 1 will not die.

## DISCUSSION

The COVID-19 pandemic presents as a public health challenge and emergency. Limited data is available in the US to guide management with effective treatment. To the best of our knowledge, no study has compared the outcome in Tocilizumab patients to a control group. For COVID-19 infection, clinical studies have shown that serum levels of inflammatory mediators are significantly higher in patients with severe disease. Excessive immune responses can trigger cytokine storms and cause damage to multiple target organs. Recent guidelines also point that a progressive rise in IL-6 may be a clinical warning indicator for the deterioration of COVID-19^13^. Tocilizumab, a monoclonal antibody against interleukin 6, emerged as an alternative treatment for COVID patients with a risk of cytokine storms recently^14^. In our study, we aim to discuss the treatment response of therapy in COVID 19 infected patients in terms of survival outcome.

This report provides a perspective on patients admitted with confirmed COVID-19 in both Intensive care and floor setting. Our hospital serves an ethnically and socioeconomically diverse population being close to New York City. A significant number of patients displayed abnormal laboratory measurements at the time of admission. A COVID-19 laboratory order set was used in the EMR to order labs at the time of hospital admission or ICU transfer which included IL-6, ferritin, CRP, triglyceride, Fibrinogen, D-dimer, CMP and CBC with differential. The majority of patients were admitted to the ICU because of acute hypoxemic respiratory failure that required respiratory support. Endotracheal intubation and invasive mechanical ventilation were needed in a significant proportion of the patients. Several key findings were observed. Several key findings were observed in the Tocilizumab group laboratory values of IL-6, Ferritin, Triglyceride and Fibrinogen were significantly higher indicating that these patients demonstrated cytokine storms. Early laboratory evaluation may be crucial in identifying cytokine storm, and may also aid clinicians in identifying patients at high risk of decompensation, ICU admission, and potentially even death. A study from China reported that increased expression of interleukin (IL)-2R and IL-6 in serum appears to predict the severity and prognosis of patients with COVID-19^18^. Elevated levels of the inflammatory indicator IL-6 in the blood have been reported to be predictive of a fatal outcome in patients with COVID-19.^19^

Out of all patients who received Tocilizumab 84.09 % were male and 15.91% were females. The overall survival rate was 61.36% in patients who received Tocilizumab compared to 48% in the control group. 59.46% percentage of male patients survived compared to 71.43% females if they received Tocilizumab. Total 72 males and 22 females were enrolled in this study, indicating that male gender has predominance of being affected with COVID-19. Our study should be considered in light of several limitations. In the study group 5 patients did not have fibrinogen level tested before they received Tocilizumab and in the control group 11 patients did not have the results for fibrinogen.

## CONCLUSION

In the context of the urgent need for effective therapies in the current pandemic, particularly in severe cases. However, there is no systematically recommended treatment for COVID-19. Tocilizumab is a humanized monoclonal antibody against the in-terleukin-6 receptor (IL-6R) and is FDA-approved for cytokine release syndrome and recently, has been administered experimentally in the treatment of severe COVID-19 pneumonia in China and Italy with promising results^16^. Cytokine Release Syndrome (CRS) occurs in a large number of patients with severe COVID-19, which is also an important cause of death. IL-6 is the key molecule of CRS, so Tocilizumab may be of value in improving outcomes. In our study Tocilizumab group seemed to have improved survival outcome. Results have to be interpreted with caution since this a retrospective study and mortality is affected by multiple, confounding factors. In the largest clinical trials database (clinicaltrials.gov) there are 35 ongoing studies registered around the world regarding the use of Tocilizumab in COVID-19 patients. We await the results of ongoing randomized controlled trials to definitely answer the question of whether Tocilizumab improves survival in COVID-19 ARDS patients.

## Data Availability

Data was obtained from electronic medical record review.

